# Midwives’ attitudes toward participation of pregnant women in a preventive vaccine hypothetical clinical trial

**DOI:** 10.1101/2021.05.09.21256815

**Authors:** Amandine Gagneux-Brunon, Emilie Guyot, Maëlle Detoc, Elisabeth Botelho-Nevers, Tiphaïne Raia-Barjat

**Affiliations:** Department of Infectious and Tropical Diseases, University Hospital of Saint-Etienne, Saint-Etienne, France; CIC 1408 INSERM, Vaccinology, University Hospital of Saint-Etienne, Saint-Etienne, France; Centre International de Recherche en Infectiologie, Team GIMAP, Univ Lyon, Université Jean Monnet, Université Claude Bernard Lyon 1, Inserm, U1111, CNRS, UMR530 1408, INSERM, University Hospital of Saint-Etienne, France; Department of Gynecology and Obstetrics, University Hospital of Saint-Etienne, Saint-Etienne, France; INSERM, SAINBIOSE, U1059, Université Jean-Monnet; Saint-Etienne, France

**Keywords:** pregnancy, vaccine, clinical trial, promotion, midwives

## Abstract

**Introduction:** Pregnant women are frequently excluded from clinical trials. Yet, inclusion of pregnant women is of interest in vaccinology including during health crisis. Promotion of clinical trials by midwives may facilitate the decision making of pregnant women. Attitudes of midwives about participation in a vaccine clinical trial have been little explored.

**Methods:** We conducted an anonymous survey from the 11th of September to the 11th of November 2020. Primary endpoint was the willingness to encourage pregnant women to participate in a hypothetical respiratory syncytial virus (RSV) vaccine clinical trial.

**Results:** Among 398 midwives who answered the questionnaire, 113 (28.3 %) were likely to encourage pregnant women to participate in the vaccine clinical trial, this proportion ranged from 25 % in senior midwives to 34.5 % among the students. After adjustment on age, parenthood, previous vaccine hesitancy attitudes, and the 5 components of the 5C model, the only predictor of the promotion of the clinical trial was the training score with an adjusted odds ratio of 1.09 (1.01-1.18, p=0.027) for a one-point increase. Vaccine hesitancy and psychological antecedents of vaccinations were not associated with a lower promotion of pregnant women trial participation among midwives.

**Conclusion:** Few respondents were likely to encourage pregnant women to participate in a vaccine clinical trial. Midwives who considered having a good training about vaccines were more prone to encourage pregnant women to participate in a RSV vaccine clinical trial.

**Problem or Issue:** Recruitment of pregnant women in vaccine clinical trial is challenging

**What is Already Known:** Pregnant women are more prone to accept participation in a clinical trial if the proposal is made by a midwife.

**What this Paper Adds:** Evaluation of attitudes and their determinants of midwives about vaccine clinical trial participation of pregnant women.

## Introduction

Pregnant women are frequently excluded from clinical trials [1]. Even, they may benefit from the study interventions, pregnant women are usually not eligible for clinical trials enrollment, due to potential harms for them and the fetus [2]. Consequently, most of them are exposed to medications not evaluated if administered during the pregnancy [3].

Vaccines are no exception and are rarely evaluated in pregnant women [1]. Immunization of pregnant women have been shown of high interest [4]. First, vaccine preventable diseases like seasonal influenza and COVID-19 are associated with an increased morbidity in pregnant women [4– 6]. Secondly, it is necessary to develop vaccines against pathogens causing congenital and perinatal infections [7]. Then, mothers may also represent the potential source of severe infections in newborns, as Respiratory Syncytial Virus (RSV) infections and pertussis. Mother immunization may lead to reduce the risk of infection in newborns by reducing pathogens carriage in mothers, and by passive transfer of antibodies [4].

Since few years, physicians and scientists call for inclusion of pregnant women in clinical trials, particularly in the context of the Zika virus epidemic and COVID-19 pandemic [2,8]. Intentions to take part in a vaccine clinical trial in pregnant women were little explored in general, safety concerns for the baby were shown to be the primary barrier to participation [9]. It has been shown that a healthcare provider recommendation may encourage pregnant women to take part in a vaccine clinical trial [10]. Midwives recommend routine immunization in women during pregnancy or in the post-partum period. Their attitude toward the participation of pregnant women in vaccine clinical trial is under explored. Our aims were to determine attitudes of midwives and midwifery students toward participation of pregnant women in a vaccine clinical trial, and to identify factors associated with the recommendation to participate.

## Participants, Ethics and Methods

We conducted an anonymous online survey (Lime Survey®) between the 11^th^ of September and the 11^th^ of November 2020. A web-link was sent by e-mail to midwives by professional organizations and by schools to midwifery students in the Auvergne-Rhône-Alpes area (France). This study was approved by the institutional review board IRBN1192020/CHUSTE.

The questionnaire collected among midwifes and midwifery students : (1) demographical characteristics, (2) vaccine hesitancy, (3) psychological antecedents of vaccinations (5C-model) [11], (4) attitudes toward routine vaccination during pregnancy, (5) perceptions about vaccines training (6) attitudes toward a hypothetical vaccine clinical trial evaluating efficacy and safety of a vaccine against RSV infections in pregnant women. A hypothetical scenario was proposed: “The research team in vaccinology of our region is conducting a clinical trial of a RSV vaccine during pregnancy, they asked you to promote the study to your patients, will you accept to promote the clinical trial?” We focused on a RSV vaccine clinical trial because RSV is an infection with vaccines currently in development for maternofetal immunization. We evaluated participants’ self-reported vaccine hesitancy according to the WHO definition using three previously adapted questions: “Have you ever refused a vaccine for yourself or a child because you considered it as useless or dangerous?” “Have you ever postponed a vaccine recommended by a physician because of doubts about it?” “Have you ever had a vaccine for a child or yourself despite doubts about its efficacy” [14]. If a participant answered yes to one of these proposals, he or she was considered to be “vaccine hesitant”. For the 5C Scale, we used the long version adapted in French and computed mean score for confidence, complacency, constraints, calculation and collective responsibility [11]. To evaluate perceptions of training about vaccines, we used 4 items with a 5-point Lickert scale “you are sufficiently trained about vaccination”, “you feel comfortable when you answer patients’ question about vaccines”, “you have access to tools to recommend vaccines to pregnant women.”, “Continuous education about vaccines is useful.” We computed a 20-point training score recoding Lickert: “totally agree=5, Slightly agree =4, do not know=3, Slightly disagree=2, Totally disagree=1”

We ran ordinal regression models to examine predictive demographic factors and attitudes of respondents’ willingness to recommend participation in the clinical trial of their pregnant patients. To identify suitable candidate variables for regression models, we first conducted univariate analysis using a chi-squared test. Variables with a p value< 0.2 in univariate analyses were then included in a multivariable regression model. Data were analyzed using SPSS version 24.0.

## Results

Questionnaire was sent to 2.618 midwives, and to 537 midwifery students. Three hundred ninety-eight answers were received; the global response rate was 12.6 %, ranging from 9.8 % in midwives and 26.4 % in students. Demographic characteristics are depicted in Table 1. Previous attitudes of vaccine hesitancy were identified in 149 (37.4%) of the responders. Mean and standard deviations for confidence, complacency, constraints, calculation, and collective responsibility scores were 3.8 (0.76), 1.49 (0.62), 1.35 (0.55), 3.8 (0.92), 4.7 (0.52) respectively.

**Table 1.**
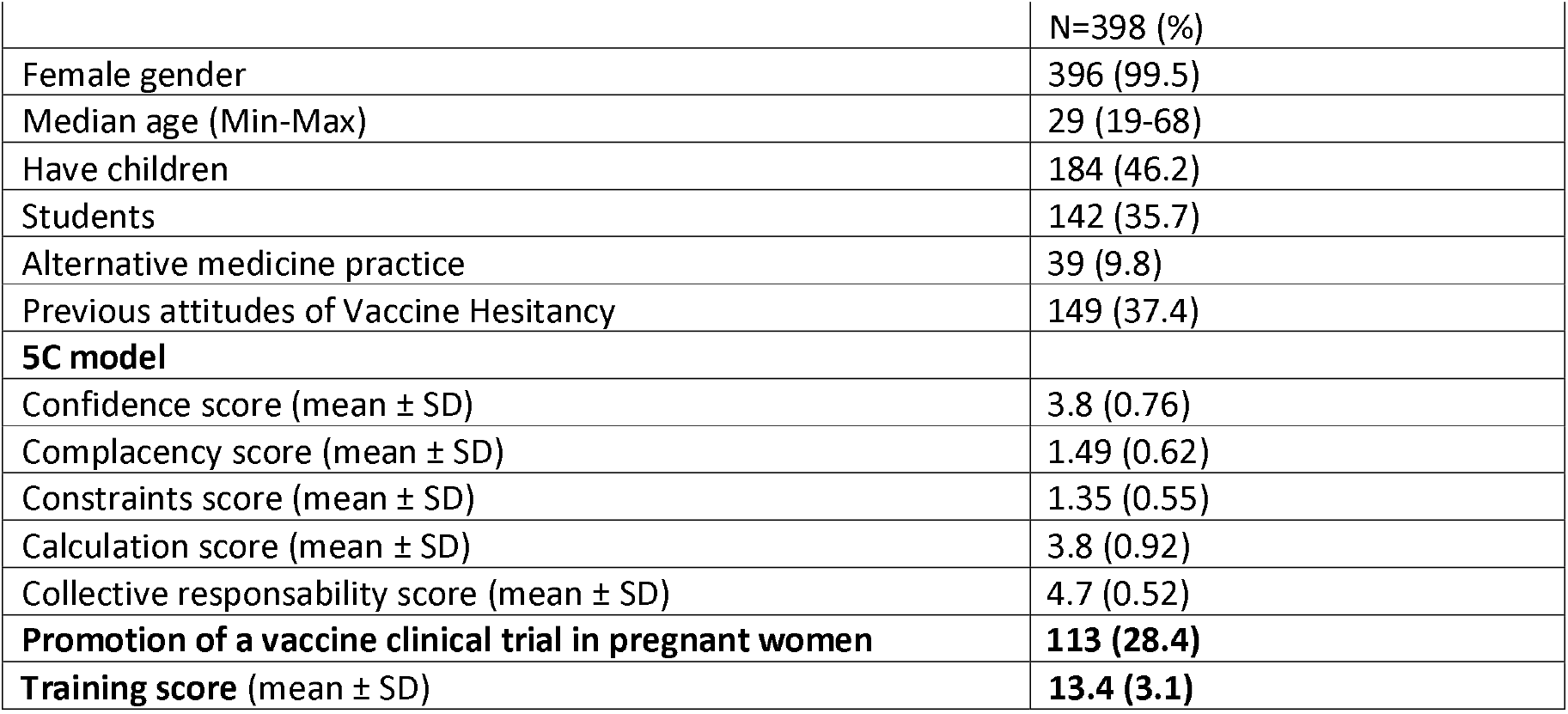
Demographic characteristics of the respondents. (Min: Minimum, Max:Maximum, SD: standard deviation)

Among the 398 responders, 113 (28.4 %) were likely to encourage participation of pregnant women in the trial. Factors associated with the promotion of the clinical trial are presented in Table 2. Midwifery students (34.5 %) were more prone than midwives (25 %) to encourage participation in the clinical trial in univariate analysis. After adjustment on age, parenthood, previous vaccine hesitancy attitudes, and the components of the 5C model, the only predictor of the promotion of the vaccine clinical trial was the training score. One-point increase in the training score was associated with an adjusted odds ratio for the promotion of the clinical trial in pregnant women of 1.09 (1.01-1.18, p=0.027) for a one-point increase. No 5C psychological antecedents of vaccinations were associated with the promotion of a vaccine clinical trial toward pregnant women by midwives and midwifery students.

**Table 2 :** Factors associated with the promotion of a vaccine clinical trial against respiratory syncitial virus in pregnant women (Min: Minimum, Max:Maximum, SD: standard deviation, aOR: adjusted odds ratio, aOR are expressed with 95 % Confidence interval)

|  | Bivariate analysis |  |  | Multivariable analysis |  |
| --- | --- | --- | --- | --- | --- |
|  | Promoting Vaccine clinical trial (113) | Not promoting vaccine clinical trial (285) | p | aOR (95 % CI) | P |
| Female gender | 112 (99.6) | 284 (99.1°) | 0.5 | NA | NA |
| Mean age | 31.0 (12.4) | 33.7 (12.3) | 0.053 | 1.0 (0.97-1.04) | 0.94 |
| Have children | 42 (37.2) | 142 (49.8) | 0.02 | 0.76 (0.34-1.71) | 0.51 |
| Students | 49 (43.4) | 93 (32.6) | 0.04 | 0.7 (0.36-1.34) | 0.28 |
| Alternative medicine practice | 8 (7.1) | 31 (10.9) | 0.25 |  |  |
| Previous attitudes of Vaccine Hesitancy | 29 (25.7) | 120 (42.5) | 0.002 | 0.6 (0.35-1.02) | 0.06 |
| **5C model** |  |  |  |  |  |
| Confidence score (mean ± SD) | 4.0 (0.72) | 3.73 (0.76) | 0.002 | 1.22 (0.83-1.79) | 0.31 |
| Complacency score (mean ± SD) | 1.4 (0.52) | 1.52 (0.64) | 0.08 | 1.05 (0.67-1.64) | 0.84 |
| Constraints score (mean ± SD) | 1.27 (0.5) | 1.37 (0.56) | 0.13 | 0.77 (0.48-1.25) | 0.77 |
| Calculation score (mean ± SD) | 3.63 (1) | 3.87 (0.88) | 0.017 | 0.86 (0.67-1.11) | 0.25 |
| Collective responsibility score (mean ± SD) | 4.82 (0.38) | 4.69 (0.56) | 0.024 | 1.16 (0.65-2.05) | 0.62 |
| **Training score (mean ± SD)** | **14 (2.9)** | **13.1 (3.1)** | **0.009** | **1.09 (1.01-1.18)** | **0.027** |

## Discussion

Participation of pregnant women in clinical trials is crucial to evaluate safety of efficacy of vaccines in this population. The attitudes of pregnant women toward participation in a vaccine clinical trial were little studied. While a great majority of pregnant women considered that is necessary to obtain safety data during pregnancy for medications, only a minority would consider participation in a clinical trial [9]. The role of midwives to encourage pregnant women participation in a clinical trial is key. For pregnant women, the best method of being asked for participation in a vaccine clinical trial was reported to be face-to-face discussion with their midwife, showing the crucial role of midwifes to promote participation in vaccine trials [12]. We reported here for the first time, the attitudes of midwives about pregnant women participation in a vaccine clinical trial in France. In our study carried out in Auvergne Rhône-Alpes region in France (the second most populated region in France with around 3.000 midwives and midwifery students), few midwives (28.4 %) were likely to promote the participation of pregnant women in a hypothetical RSV vaccine clinical trial. In a study carried out in England, this proportion was lower than 68 % of midwives encouraging pregnant women to participate in a RSV vaccine clinical trial [12]. Midwives familiar with RSV infections were more likely to encourage participation in the vaccine clinical trial, data not explored in our work [12]. Wilcox et al. also evaluated the attitudes of obstetricians, and 92 % of them were likely to encourage pregnant women participation in a RSV vaccine clinical trial [12]. However, in contrast with Wilcox et al., we investigated the impact of midwives’ vaccine hesitancy and/or psychological antecedents of vaccination on their attitudes toward participation of pregnant women in a vaccine clinical trial. Vaccine hesitancy of healthcare worker may impact their attitudes toward vaccine clinical trials participation of their patients [13]. In spite of a great proportion of midwives or midwifery students with previous attitudes of vaccine hesitancy, we did not observe any impact in multivariable analysis of vaccine hesitancy or of the psychological antecedents of vaccinations (onfidence, complacency, constraints, calculation and collective responsibility) on the attitude of promotion of a vaccine clinical trial to pregnant women. To our knowledge, our study is the first study using the 5C scale adapted in French. The validation of the 5C scale adapted in French (after translation and back translation) was undergoing during the study period [11].

We observed here that specific training about vaccines was associated with attitude to recommend participation in the hypothetical vaccine clinical trial. We can also suggest that specific training about clinical research will empower midwives to approach pregnant women for participation in a clinical trial. We previously observed that primary care physicians with an experience in clinical research were more prone to participate in the decision making of their patients approached for participation in a vaccine clinical trial [13]. To optimize recruitment and retention of pregnant women in a vaccine clinical trial, specific training about the vaccine preventable disease, vaccines, and clinical research of midwives might particularly be of interest and effective [14].

Our study suffers from several limitations. First, the promotion of a vaccine clinical trial was one part of a survey about practices of vaccination in midwives, consequently, we did not evaluate knowledge, attitudes and perception of midwives about clinical research. We previously observed that knowledge about clinical research was crucial to help primary care physicians to encourage their patient for clinical trial participation [13]. Knowledge, attitudes and perceptions of clinical research need to be evaluated in midwives. Secondly, we only considered the promotion of a hypothetical RSV vaccine, whereas, at this time France was facing the COVID-19 pandemic second wave. Due to the emergency, it is possible that midwives will encourage participation of pregnant women in a COVID-19 vaccine trial. It is uncertain that attitudes may not be associated with the different type of vaccines or of diseases. Main concerns about participation in a vaccine clinical trial for pregnant women focused on safety for the baby, a RSV vaccine will protect the baby if effective. As a COVID-19 vaccine administered to a pregnant woman will mainly protect the women from severe COVID-19, it is uncertain that midwives will be more prone to promote pregnant participation in a COVID-19 vaccine clinical trial.

In conclusion, few midwives were little prone to promote vaccine clinical trial participation of pregnant women while their role is crucial. The main predictor of the attitude of promotion of vaccine clinical trial is the knowledge and the training about vaccines in general. The improvement of the training about clinical research, vaccines, vaccine preventable diseases, may probably increase the proportion of midwives encouraging pregnant women to participate in a vaccine clinical trial.

**Authors have no competing interest with this work**.

## Data Availability

Data are available after contact with the corresponding author

## References

1. Beigi RH, Krubiner C, Jamieson DJ, Lyerly AD, Hughes B, Riley L, Faden R, Karron R. The need for inclusion of pregnant women in COVID-19 vaccine trials. Vaccine. févr 2021;39(6):868⍰70.

2. Heyrana K, Byers HM, Stratton P. Increasing the Participation of Pregnant Women in Clinical Trials. JAMA. 27 nov 2018;320(20):2077⍰8.

3. Ayad M, Costantine MM. Epidemiology of medications use in pregnancy. Semin Perinatol. 1 nov 2015;39(7):508⍰11.

4. Loubet P, Anselem O, Launay O. Immunization during pregnancy. Expert Rev Vaccines. 2018;17(5):383⍰93.

5. Lokken EM, Huebner EM, Taylor GG, Hendrickson S, Vanderhoeven J, Kachikis A, Coler B, Walker CL, Sheng JS, al- Haddad Bjs, McCartney SA, Kretzer NM, Resnick R, Barnhart N, Schulte V, Bergam B, Ma KK, Albright C, Larios V, Kelley L, Larios V, Emhoff S, Rah J, Retzlaff K, Thomas C, Paek BW, Hsu RJ, Erickson A, Chang A, Mitchell T, Hwang JK, Erickson S, Delaney S, Archabald K, Kline CR, LaCourse SM, Adams Waldorf KM. Disease Severity, Pregnancy Outcomes and Maternal Deaths among Pregnant Patients with SARS-CoV-2 Infection in Washington State. Am J Obstet Gynecol [Internet]. 27 janv 2021 [cité 1 févr 2021]; Disponible sur: http://www.sciencedirect.com/science/article/pii/S0002937821000338

6. Vousden N, Bunch K, Knight M, UKOSS Influenza Co-Investigators Group. Incidence, risk factors and impact of seasonal influenza in pregnancy: A national cohort study. PloS One. 2021;16(1):e0244986.

7. Singh T, Otero CE, Li K, Valencia SM, Nelson AN, Permar SR. Vaccines for Perinatal and Congenital Infections—How Close Are We? Front Pediatr [Internet]. 2020 [cité 12 mars 2021];8. Disponible sur: https://www.frontiersin.org/articles/10.3389/fped.2020.00569/full

8. Minkoff H, Ecker J. Balancing risks: making decisions for maternal treatment without data on fetal safety. Am J Obstet Gynecol. 1 févr 2021;

9. Palmer S, Pudwell J, Smith GN, Reid RL. Optimizing Participation of Pregnant Women in Clinical Trials: Factors Influencing Decisions About Participation in Medication and Vaccine Trials. J Obstet Gynaecol Can. 1 oct 2016;38(10):945⍰54.

10. Goldfarb IT, Jaffe E, James K, Lyerly AD. Pregnant women’s attitudes toward Zika virus vaccine trial participation. Vaccine. 29 oct 2018;36(45):6711⍰7.

11. Betsch C, Bach Habersaat K, Deshevoi S, Heinemeier D, Briko N, Kostenko N, Kocik J, Böhm R, Zettler I, Wiysonge CS, Dubé É, Gagneur A, Botelho-Nevers E, Gagneux-Brunon A, Sivelä J. Sample study protocol for adapting and translating the 5C scale to assess the psychological antecedents of vaccination. BMJ Open. mars 2020;10(3):e034869.

12. Wilcox CR, Calvert A, Metz J, Kilich E, MacLeod R, Beadon K, Heath PT, Khalil A, Finn A, Snape MD, Vandrevala T, Nadarzynski T, Coleman MA, Jones CE. Attitudes of Pregnant Women and Healthcare Professionals Toward Clinical Trials and Routine Implementation of Antenatal Vaccination Against Respiratory Syncytial Virus: A Multicenter Questionnaire Study. Pediatr Infect Dis J. sept 2019;38(9):944⍰51.

13. Detoc M, Touche C, Charles R, Lucht F, Gagneux-Brunon A, Botelho-Nevers E. Primary physicians’ attitudes toward their patients receiving a proposal to participate in a vaccine trial. Hum Vaccines Immunother. 26 juin 2019;11⍰1.

14. Frew PM, Saint-Victor DS, Isaacs MB, Kim S, Swamy GK, Sheffield JS, Edwards KM, Villafana T, Kamagate O, Ault K. Recruitment and Retention of Pregnant Women Into Clinical Research Trials: An Overview of Challenges, Facilitators, and Best Practices. Clin Infect Dis. 15 déc 2014;59(suppl_7):S400⍰7.

